# Global, regional, and national estimates of tuberculosis incidence averted by eliminating undernutrition in adults: a modelling study

**DOI:** 10.1101/2025.08.21.25334062

**Authors:** Matthew J Saunders, C Finn McQuaid, Pranay Sinha, Leonardo Martinez, James A Seddon, Peter J Dodd

## Abstract

**Background:** Current efforts to reduce global tuberculosis incidence have proved insufficient, highlighting that urgent action is needed to address underlying modifiable risk factors such as undernutrition. We aimed to estimate the global impact of eliminating undernutrition on tuberculosis incidence among adults accounting for varying nutritional status by country, sex, and age, in addition to incorporating the continuous, non-linear relationship between body mass index (BMI) and tuberculosis risk.

**Methods:** We used a continuous risk framework to consider the population-level implications of BMI distributions for tuberculosis incidence for those aged ≥15 years. We generated BMI distributions for each country, sex, and age group applying a bilinear model for the logarithmic relative risk of tuberculosis incidence at different BMI values. We assessed the impact of eliminating moderate/severe undernutrition (BMI<17kg/m^2^) or all undernutrition (BMI<18.5kg/m^2^) on tuberculosis incidence by constructing counterfactual BMI distributions that redistributed those with low BMI to higher BMI, proportional to the remaining density.

**Findings:** We estimated that eliminating moderate/severe undernutrition would avert 1.4 million (95%UI, 1.1-1.7) tuberculosis episodes globally, representing 16.8% (14.3-19.2) of global adult incidence, while eliminating all undernutrition would avert 2.3 million (1.8-2.7) episodes, a reduction of 26.5% (23.2-29.8). The largest proportional reductions in tuberculosis incidence could be achieved by eliminating undernutrition in the African, South-East Asian, and Eastern Mediterranean regions; females; and adolescent or elderly adults.

**Interpretation:** Over a quarter of global tuberculosis incidence in adults would be averted by eliminating undernutrition, approximately three times higher than current estimates. These findings highlight the urgent need to scale up population-level nutritional interventions, which may have myriad social and health benefits beyond tuberculosis, alongside research to determine optimal implementation strategies and impacts.

**Funding:** No specific funding

## Introduction

Tuberculosis is the world’s leading cause of death from a single infection, affecting an estimated 10.8 million people in 2023, and killing 1.25 million.^1^ Global tuberculosis incidence declined by 8.3% between 2015 and 2023, far short of World Health Organization (WHO) ‘End TB Strategy’ targets of 50% by 2025 and 90% by 2035.^1,2^ Although the pace of decline has been partly slowed by the impact of COVID-19,^3^ clearly these targets are unlikely to be achieved without substantial changes in the epidemic, and in our response.^4^ While it remains important to improve the effectiveness and coverage of the biomedical interventions underpinning tuberculosis prevention, diagnosis, and treatment, it is also critical to recognise that the global tuberculosis epidemic is driven by multiple modifiable, socially-determined risk factors that could be addressed to reduce tuberculosis incidence.^5,6^

Undernutrition, which encompasses multiple forms of nutrient deficiency, but is commonly defined in adults by an underweight body mass index (BMI<18.5kg/m^2^), is one such risk factor. Having undernutrition increases both the risk of developing tuberculosis, and the risk of adverse outcomes once diagnosed and treated.^7^ Importantly, undernutrition has been shown to be a highly modifiable risk factor both historically and in contemporary contexts. Improved nutrition was a major contributor to the decline in tuberculosis incidence observed in high-income countries during the 20th century;^8^ and the recent RATIONS study in India demonstrated that providing nutritional support to members of tuberculosis-affected households could reduce tuberculosis incidence by approximately 40%.^9^ Subsequent modelling estimated a meaningful population-level impact of this intervention on tuberculosis incidence if scaled up to other tuberculosis-affected households nationally, and showed it would be highly likely to be cost-effective.^10^ Given rising food insecurity globally,^11^ there is a clear need to explore the potential of expanding nutritional interventions beyond tuberculosis-affected households to broader populations with high undernutrition prevalence.

To inform health programming, budgeting, and intervention prioritisation, it is important to understand how much tuberculosis incidence could be averted by eliminating undernutrition. To facilitate this, the WHO calculates annual estimates of the number of tuberculosis episodes caused by undernutrition (alongside other risk factors) for each country, based on population attributable fractions (PAF).^1^ The most recent global PAF estimate of 8.9% was informed by a recent Cochrane review, which estimated the relative risk of tuberculosis in a binary comparison of people with undernutrition (BMI<18.5kg/m^2^) versus those without undernutrition (BMI≥18.5kg/m^2^).^12^ However, because of the established dose-response relationship between BMI and tuberculosis risk, this binary conceptualisation of a naturally continuous exposure likely underestimates the relative risk and hence the PAF.^13,14^ Furthermore, given that BMI distributions vary by age, sex, and between and within regions, more granular estimates would enable more targeted and effective policy decisions.

A recent systematic review and dose-response meta-analysis has provided updated estimates of how tuberculosis risk changes across the entire BMI range in adults.^15^ This review showed that tuberculosis risk is highest at the lowest BMI values and decreases non-linearly as BMI increases, with the steepest reductions in tuberculosis risk occurring when moving through underweight and normal weight ranges, and more modest reductions continuing through overweight and obese ranges. In this study, we combined these new estimates with population-specific BMI distributions to model how much tuberculosis incidence in adults could be averted by eliminating undernutrition across different demographic groups and regions.

## Methods

We used a continuous risk framework to consider the population-level implications of BMI distributions for tuberculosis incidence by country, sex, and age group for those aged 15 years or older in 2023. An overview of the methods is shown in Figure 1. We focussed on adults because different anthropometric indicators are used in children, and there is limited evidence available on how these indicators influence tuberculosis risk. Ethical approval was not required as this study used only published, aggregate data.

**Figure 1:**
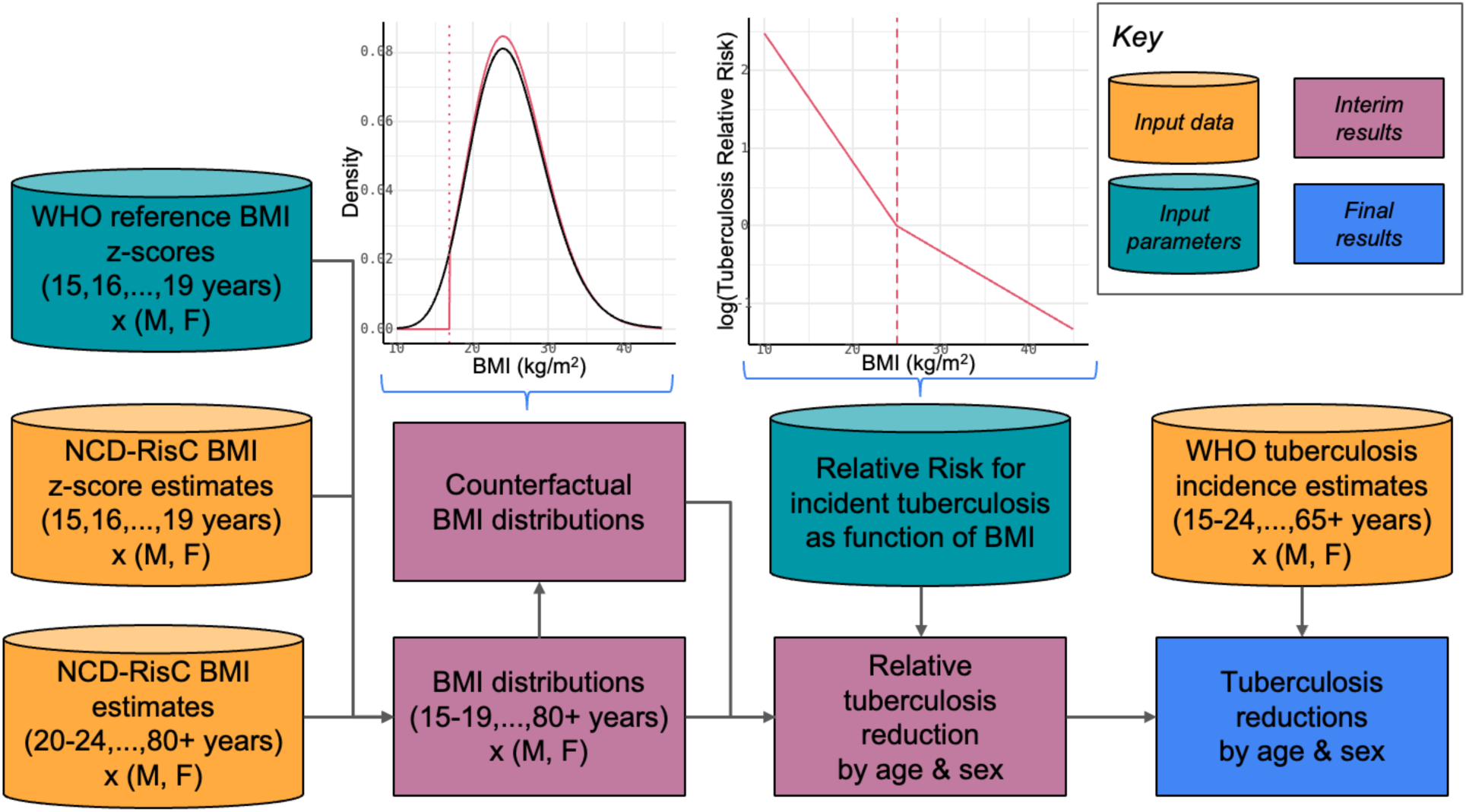
Data inputs and modelling approach. Cylinders represent input data; boxes represent modelling steps and outputs. BMI = body mass index; WHO = World Health Organization. The left graph shows an example population BMI distribution (black), and the counterfactual BMI distribution from reallocating those with BMI<17 kg/m^2^ proportionally across the rest of the distribution. The right graph shows the log-bilinear tuberculosis risk function from Saunders et al.^15^ applied to BMI distributions to calculate changes in tuberculosis incidence.

### BMI distributions

We used estimates from the NCD Risk Factor Collaboration (NCD-RisC) for their most recent year (2022) to fit BMI distributions for each country, sex, and 5-year age group.^16^ We chose gamma distributions based on computational advantages and comparable fit with log-normal distributions on an individual-level BMI test dataset from Peru (Appendix p4).

We fitted gamma distributions to their adult (20+ years) estimates by minimizing a sum-of-squares loss function that measured the difference between the population fraction in each of eight BMI ranges, for each country, sex, and age group (Appendix pp5-6), weighted by their precision. Where fitting did not converge, we assigned the distribution for the same sex and country in the age group below.

For adolescent (15-19 years) estimates, we used WHO reference BMI tables to convert the z-scores used to define reported categories into absolute BMI values, using the midpoint of the single year age categories in the reference tables. We then minimized a sum-of-squares loss function that measured the difference between the population fraction in each of five BMI ranges, for each country, sex, and single-year age group (Appendix p5), weighted by their precision. Finally, we constructed an average gamma distribution for the 15-19 year age group to match the means of the single-year gamma distribution means and variances. Where fits of single-year BMI distributions did not converge, these were dropped from this averaging procedure.

Normal approximations to the loss function around the optima were used to represent uncertainty in BMI distribution parameters as bivariate normal distributions.

### Relative risks of tuberculosis and counterfactual comparisons

We used a bilinear model for the logarithmic relative risk of tuberculosis incidence at different BMI values, based on the recent systematic review and dose-response meta-analysis of Saunders et al. (see right graph in Figure 1).^15^ Relative risks of tuberculosis incidence comparing populations were computed by taking the ratio of mean relative risks over each population’s BMI distribution, which could be calculated analytically (Appendix pp6-7).

Since there is no natural reference BMI distribution to make comparisons against, we assessed the impact of eliminating low BMI on tuberculosis incidence by constructing counterfactual BMI distributions that redistributed those with low BMI in each country, sex, and age-group to higher BMI, proportional to the remaining density. Specifically, we considered the effect of eliminating moderate/severe undernutrition (BMI<17kg/m^2^) or all undernutrition (BMI<18.5kg/m^2^). The left graph in Figure 1 illustrates the construction of a counterfactual BMI distribution (red) by eliminating moderate/severe undernutrition in this manner from a base case distribution (black). The relative risk of tuberculosis incidence for the counterfactual compared to base case distributions could again be calculated analytically (Appendix p7). Uncertainty in the slopes for the bilinear risk function was represented as a bivariate normal distribution based on variance-covariance matrix from the regression of Saunders et al.^15^

### Tuberculosis incidence and metrics calculated

To calculate the impact on tuberculosis incidence, we merged our estimates of relative reductions in each country, sex, and age group, with WHO estimates of tuberculosis incidence by country, sex, and age in 2023. Age groups used by WHO in their estimates are more aggregated than ours, and by default we assumed tuberculosis incidence was uniformly distributed over age sub-groups. Some countries provide tuberculosis notifications for the age 15-24 year groups separately. In these countries, sex-specific notification ratios were used to disaggregate 15-24 year incidence into 15-19 and 20-24 year age groups. For the WHO 65+ year age group, we used United Nations demographic estimates to split tuberculosis incidence proportional to population across 65-69, 70-74, 75-79, 80-84, and 85+ year age groups. For all estimates of tuberculosis incidence averted, we propagated uncertainty in incidence estimates analytically and combined it with uncertainty in relative risks and BMI distributions from 1,000 samples using the law of total variance.

We computed the total tuberculosis incidence - and also proportion of tuberculosis incidence in each group - averted by eliminating moderate/severe undernutrition or all undernutrition, globally and by WHO region, for each sex and both sexes combined. We computed the proportion of tuberculosis incidence averted in each age group by eliminating moderate/severe undernutrition or all undernutrition, globally and by WHO region, for each sex. Finally, we also calculated the proportion of tuberculosis incidence averted overall in each country by eliminating moderate/severe undernutrition or all undernutrition.

## Results

### BMI distributions

We obtained BMI distributions for 15 age groups, each sex, and 199 countries, representing >99% of the world population aged 15 years or older and >99% of estimated global tuberculosis incidence in this group. Of the 7600 country-sex-age groups that we fitted BMI distributions to, 7 failed to converge and were dealt with as described above.

We estimated that, among those aged 15 years or older globally, 292 million (95% uncertainty interval[UI], 285 million to 300 million) had moderate/severe undernutrition and 615 million (95%UI, 607 million to 624 million) had undernutrition, representing 4.8% (95%UI, 4.7% to 5.0%) and 10.2% (95%UI, 10.1% to 10.3%) of this group respectively (Appendix Tables A2-3). The median for the prevalence of moderate/severe undernutrition across countries was 2.3% (interquartile range, 1.3% to 5.4%), and was 5.3% (interquartile range, 3.4% to 11.9%) for the prevalence of all undernutrition. The age distribution of the prevalence of both measures of undernutrition were U-shaped across all regions and both sexes (Appendix Figure A5). Prevalence of undernutrition globally was similar between sexes (a male:female ratio of 0.99 (95%UI, 0.94 to 0.99) for moderate/severe undernutrition), but varied by WHO region. The male:female ratio for moderate/severe undernutrition was close to one in South-East Asia (0.99 (0.91 to 1)), higher than one (i.e., more undernutrition in men than women) in the African [1.42 (95%UI, 1.23 to 1.35)] and Eastern-Mediterranean [1.23 (95%UI, 1.08 to 1.1)] regions, and below one elsewhere: 0.45 (95%UI, 0.42 to 0.51) in Europe, 0.8 (95%UI, 0.74 to 0.84) in the Americas, and 0.61 (95%UI, 0.57 to 0.64) in the Western Pacific regions.

### Global and regional incident tuberculosis averted

We estimated that eliminating moderate/severe undernutrition would avert 1.4 million (95%UI, 1.1 million to 1.7 million) tuberculosis episodes globally, and that eliminating all undernutrition would avert 2.3 million (95%UI, 1.8 million to 2.7 million) tuberculosis episodes globally (Table 1). This represents a 16.8% (95%UI, 14.3% to 19.2%) reduction in adult global tuberculosis incidence for eliminating moderate/severe undernutrition and a 26.5% (95%UI, 23.2% to 29.8%) reduction in global tuberculosis incidence for eliminating all undernutrition (Figure 2). The biggest decrease in tuberculosis incidence by eliminating all undernutrition would be seen for the South-East Asian region at 1.1 million (95%UI, 0.9 million to 1.4 million), followed by the African and Western Pacific regions (Table 1). Proportional reductions in tuberculosis incidence for eliminating all undernutrition would be largest in the South-East Asian [28.8% (95%UI, 25.2% to 32.3%)], African [28.4% (95%UI, 24.9% to 32.0%)], and Eastern Mediterranean regions [27.8% (95%UI, 22.1% to 33.4%); and lower in the Western Pacific [18.9% (95%UI, 14.8% to 23.0%)], the American [13.5% (95%UI, 11.1% to 15.9%)], and European [9.8% (95%UI, 7.9% to 11.8%)] regions.

**Figure 2:**
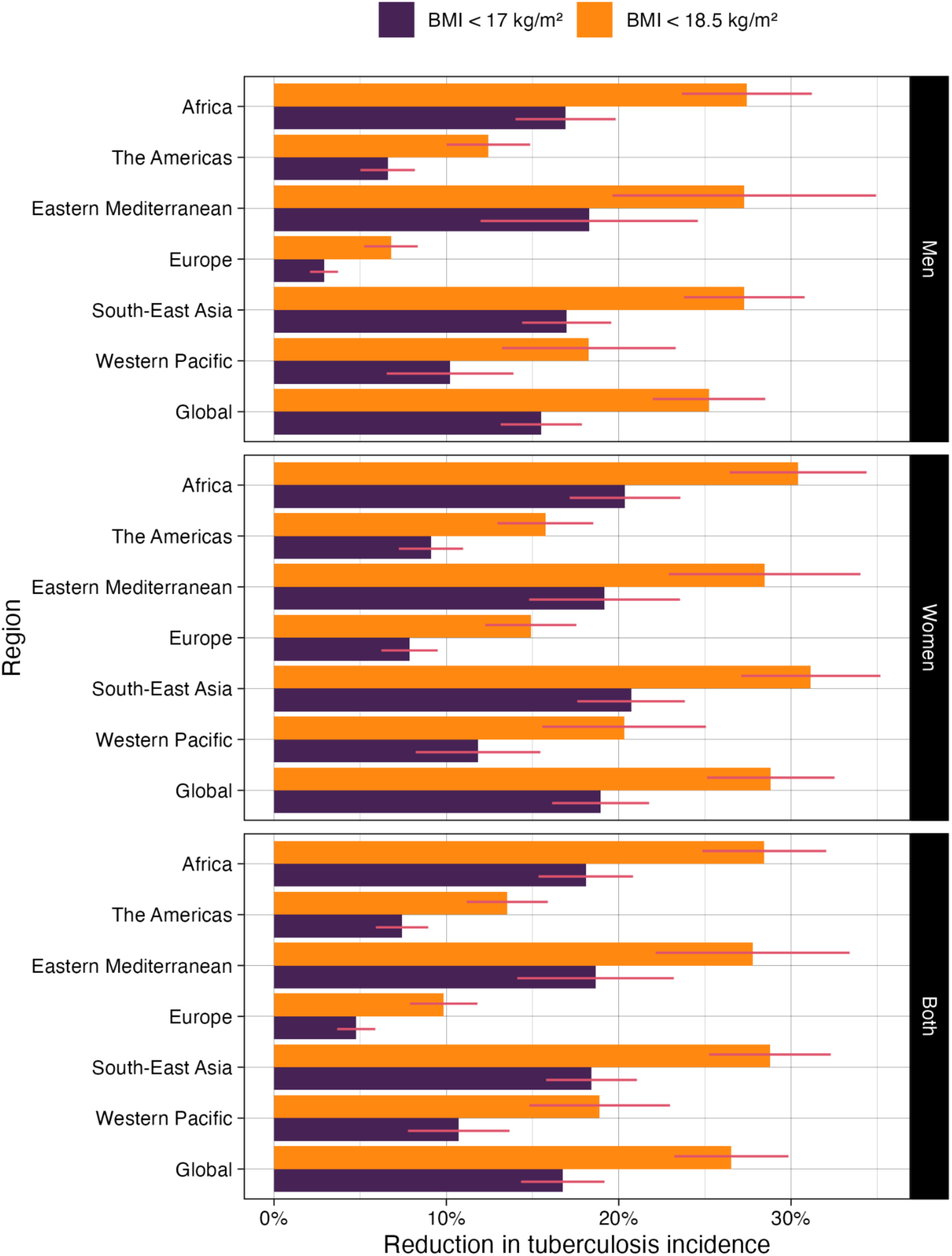
Global and regional proportion of tuberculosis incidence for 2023 in men and women averted by eliminating moderate/severe undernutrition (body mass index [BMI] < 17kg/m^2^) or all undernutrition (BMI < 18.5kg/m^2^).

**Table 1:** Global and regional tuberculosis incidence for 2023 in men and women averted by eliminating moderate/severe undernutrition (body mass index [BMI] < 17kg/m^2^) or all undernutrition (BMI < 18.5kg/m^2^).

|  | **Moderate/severe undernutrition**<br>**BMI < 17kg/m<sup>2</sup>** |  |  | **All undernutrition**<br>**BMI < 18.5kg/m<sup>2</sup>** |  |  |
| --- | --- | --- | --- | --- | --- | --- |
| **Region** | Men | Women | **Both** | Men | Women | **Both** |
| Africa | 217,000 (132,000 to 302,000) | 160,000 (99,700 to 220,000) | 377,000 (268,000 to 486,000) | 357,000 (245,000 to 469,000) | 243,000 (167,000 to 320,000) | 600,000 (457,000 to 743,000) |
| The Americas | 9,960 (4,680 to 15,200) | 7,800 (4,270 to 11,300) | 17,800 (11,100 to 24,400) | 20,200 (12,200 to 28,200) | 14,100 (9,080 to 19,100) | 34,300 (24,300 to 44,300) |
| Eastern Mediterranean | 65,000 (3,990 to 126,000) | 57,100 (7,230 to 107,000) | 122,000 (42,400 to 202,000) | 103,000 (26,300 to 179,000) | 87,100 (25,400 to 149,000) | 190,000 (90,300 to 289,000) |
| Europe | 2,910 (0 to 6,450) | 4,170 (1,340 to 7,000) | 7,080 (2,720 to 11,400) | 7,350 (1,720 to 13,000) | 8,170 (4,160 to 12,200) | 15,500 (8,430 to 22,600) |
| South-East Asia | 405,000 (239,000 to 570,000) | 309,000 (174,000 to 445,000) | 714,000 (491,000 to 937,000) | 666,000 (451,000 to 880,000) | 474,000 (307,000 to 641,000) | 1,140,000 (854,000 to 1,430,000) |
| Western Pacific | 102,000 (0 to 244,000) | 53,700 (3,500 to 104,000) | 156,000 (23,300 to 288,000) | 191,000 (30,900 to 351,000) | 96,000 (30,800 to 161,000) | 287,000 (113,000 to 461,000) |
| **Global** | 802,000 (557,000 to 1,050,000) | 592,000 (416,000 to 768,000) | 1,390,000 (1,070,000 to 1,720,000) | 1,340,000 (1,020,000 to 1,670,000) | 923,000 (702,000 to 1,140,000) | 2,270,000 (1,830,000 to 2,700,000) |

### Tuberculosis averted by sex, age, and country

More tuberculosis episodes would be averted among men than women globally by eliminating all undernutrition (Table 1), in line with the larger number of global tuberculosis episodes among men. However, the proportion of tuberculosis incidence averted globally by eliminating all undernutrition would be slightly higher among women [28.8% (95%UI, 25.1% to 32.5%)] than men [25.2% (95%UI, 22.0% to 28.5%); Figure 2]. Moreover, these patterns vary by region. While the number of tuberculosis episodes averted would be higher in men than women for all regions except Europe, the proportion of tuberculosis incidence averted would be higher in women in all regions - up to twice as high in Europe (male:female ratio in proportion reduction 0.46 (95%UI, 0.32 to 0.59)).

The highest number of tuberculosis episodes and proportions of tuberculosis incidence averted by eliminating all undernutrition would be seen in African and Asian countries (Figure 3). Eliminating all undernutrition would reduce tuberculosis incidence by over 25% in 47 countries: 30 countries on the African continent, six countries in South-East Asia, and six in the Americas (Appendix Figure A6).

**Figure 3:**
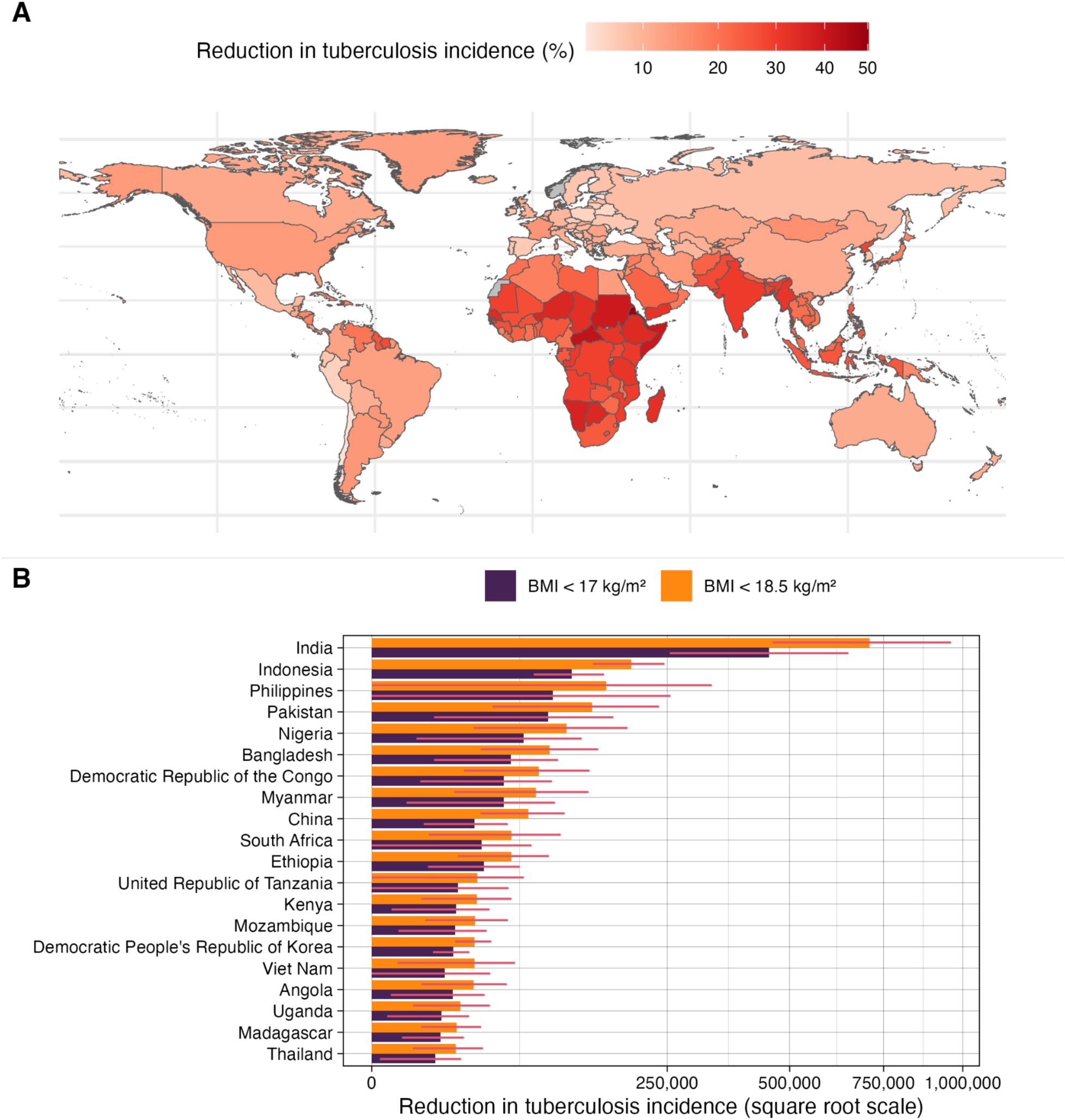
Proportional and absolute tuberculosis incidence averted for 2023 by reducing undernutrition. **A**) Map colour (square root scale) shows percentage reduction in tuberculosis incidence by country from eliminating all undernutrition (body mass index [BMI] < 18.5kg/m^2^). **B**) The countries with the highest 20 absolute reductions in tuberculosis incidence from eliminating moderate/severe undernutrition (BMI < 17kg/m^2^) and all undernutrition (BMI < 18.5kg/m^2^). Red error bars show 95% uncertainty intervals.

The proportion of tuberculosis incidence averted by eliminating all undernutrition globally showed a U-shaped dependence on age, with the highest proportions averted in adolescent and old-age populations (Figure 4). This pattern also varies by region, being larger in older age groups in the African region, and exhibiting asymmetric age patterns by sex in the African and European regions (Appendix Figure A7).

**Figure 4:**
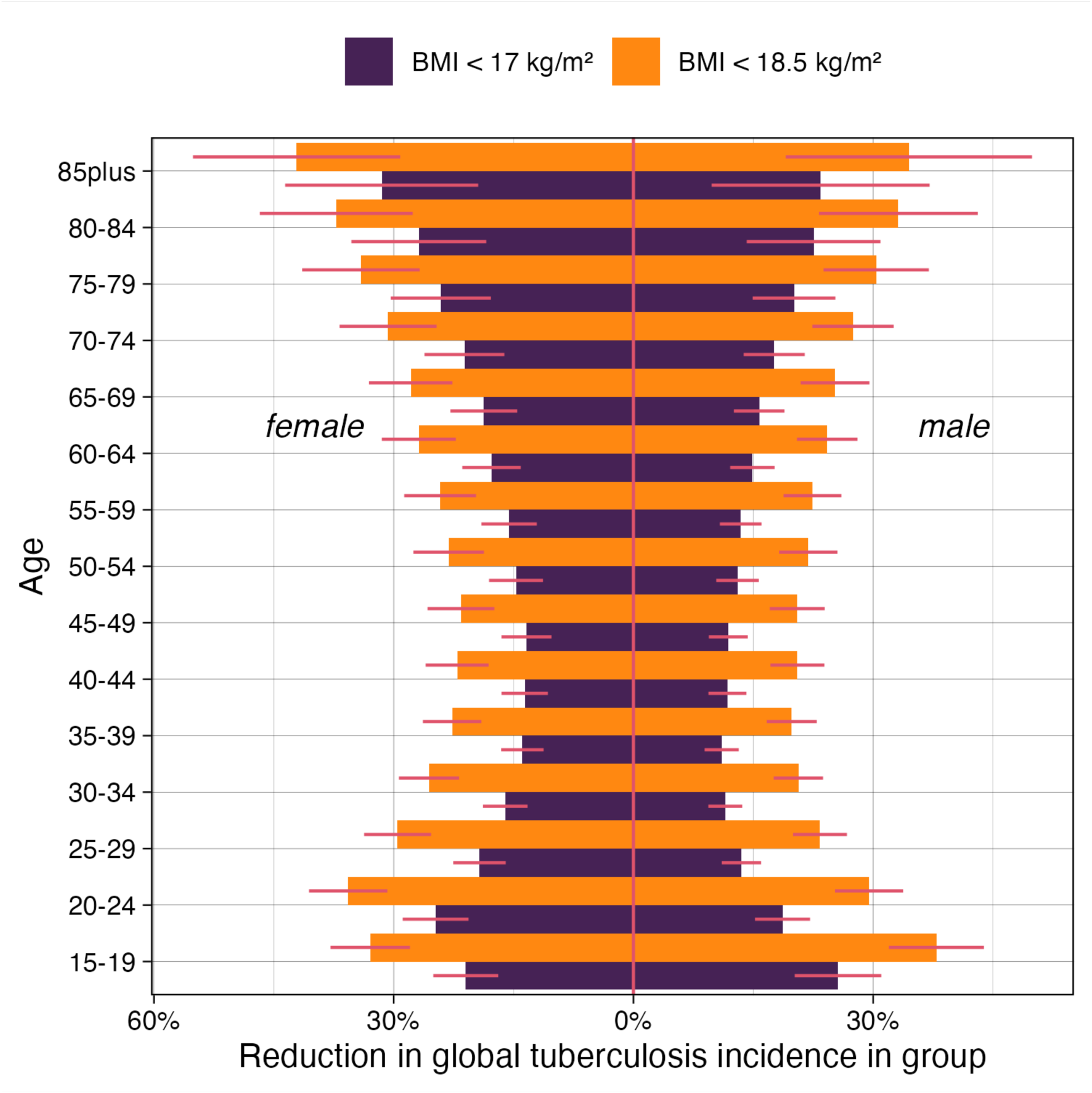
Global proportion of tuberculosis incidence for 2023 in each sex and age group averted by eliminating moderate/severe undernutrition (body mass index [BMI] < 17kg/m^2^) or all undernutrition (BMI < 18.5kg/m^2^).

## Discussion

Our modelling suggests that nearly 2.3 million tuberculosis episodes in 2023 (27% of global tuberculosis incidence in adults) would be averted if all adults with undernutrition, representing 10% of this group globally, had their BMI increased above 18.5kg/m^2^. Strikingly, nearly 1.5 million tuberculosis episodes (17% of global tuberculosis incidence in adults) in 2023 would be averted if all adults with moderate/severe undernutrition, representing just 5% of this group globally, had their BMI increased above 17kg/m^2^. The proportion of tuberculosis incidence averted by eliminating undernutrition varied both between and within regions, and by sex and age, highlighting the importance of disaggregated data for informing policy decisions and allocation of resources. Broadly, our results suggest that the largest proportional reductions in incidence could be achieved by eliminating undernutrition in the African, South-East Asian, and Eastern Mediterranean regions; females; and adolescents and elderly adults.

Our estimate that 27% of global tuberculosis incidence in adults could be averted by eliminating undernutrition, which is functionally equivalent to a PAF, is around three times higher than the most recent WHO PAF estimate of 8.9%.^1^ The WHO estimate uses a binary conceptualisation of nutritional status and risk, and a crude prevalence of undernutrition in each country to calculate the PAF, which is subject to methodological limitations. In contrast, we conducted a more refined analysis that used updated estimates for how tuberculosis risk changes non-linearly across the entire BMI range, and accounts for both the continuous nature of BMI as an exposure, and how BMI distributions differ across populations. Our estimates are similar to those of a previous study which generated revised PAF for undernutrition for 30 high tuberculosis burden countries.^17^ However, that analysis considered a limited number of countries and ages, and used an older risk estimate as input. While other modelling studies have also demonstrated the potential impact of changes in nutritional status on tuberculosis incidence to various degrees,^18–24^ none have provided comprehensive global estimates disaggregated by region, sex, and age. While we focussed on adults because of the different anthropometric indicators used in children, and lack of evidence on how nutritional status in children influences tuberculosis risk, our study highlights the need for further research in this population, given the overall importance of nutrition as a determinant of child development and mortality.^25^

These results highlight the fundamental importance of undernutrition in driving the global tuberculosis epidemic. While nutritional interventions for tuberculosis-affected households have been shown to be effective,^9,26^ their impact on the overall epidemic is likely to be limited because most *Mycobacterium tuberculosis* transmission occurs outside of these households.^10,27^ Our results demonstrate the urgent need for further research exploring the health and economic impacts of population-level nutritional interventions extending beyond tuberculosis-affected households, including how these interventions should be designed and prioritised to most effectively prevent tuberculosis. This is more salient than ever given converging global crises that threaten to worsen both undernutrition and tuberculosis simultaneously. Global food insecurity has increased substantially since 2015, particularly in areas of the world with a high tuberculosis burden, driven by the COVID-19 pandemic and increasing global conflict.^11,28^ Climate change is projected to further challenge food security, both in the short-term due to extreme weather events and disasters, and in the longer-term because of effects on agriculture and aquaculture productivity.^29^ Meanwhile, projected reductions in international donor funding through the United States Agency for International Development (USAID) and The Global Fund to Fight AIDS, Tuberculosis and Malaria are expected to be catastrophic for tuberculosis, and global health more generally.^30,31^ Against this backdrop, nutritional interventions are already being deprioritised in favour of maintaining biomedical interventions,^32^ despite the fact they are likely to have myriad benefits extending beyond tuberculosis, including lowering child mortality,^25^ improving mental health,^33^ and enhancing economic productivity.^34^

Translating these findings into effective, sustainable public health interventions requires consideration of multiple factors. Nutrition-specific interventions, including direct food assistance and correction of micronutrient deficiencies, are essential for people with moderate/severe undernutrition, where our data suggest the greatest relative impact on tuberculosis incidence could be achieved. These are often delivered through maternal and child health interventions, or school-based programmes, and can be combined with nutritional counselling aiming to promote healthy nutritional behaviours. However, sustainable reductions in undernutrition (and preventing undernutrition from occurring) will require the expansion of these programmes to encompass undernutrition among adults, and incorporation of nutrition-sensitive approaches that address underlying structural and social determinants. These may include broader social protection interventions such as cash transfers, universal health coverage initiatives, and other social policies including subsidising healthy foods. At a population-level, these nutritional interventions could be targeted towards communities and households with high social vulnerability indices. Recent evidence from Brazil found lower tuberculosis incidence and mortality among beneficiaries of a targeted cash transfer programme with conditions around nutrition surveillance.^35^ Importantly, people at risk of tuberculosis often have multiple nutritional deficiencies, with both macro- and micro-nutrient deficiencies increasing risk.^7^ Vertical programmes targeting a small subset of these have been unsuccessful in reducing tuberculosis incidence.^36^ Therefore, interventions aiming to improve overall food security and nutritional status are likely to offer the greatest promise for reducing tuberculosis incidence.

The broad scope of potential interventions raises important questions about funding responsibility, highlighting the need for more work to understand the budgetary and cost implications of scale-up. While tuberculosis-specific donors like The Global Fund have traditionally focused on biomedical interventions, addressing undernutrition at a population level requires coordination between health, development, and humanitarian funding streams, alongside stronger domestic financing and political commitment, including through innovative fund-raising mechanisms, e.g. taxes on sugary drinks or calorie-dense foods with little nutritional value. Furthermore, interventions must be designed with awareness of the double burden of malnutrition, as many settings now face coexisting undernutrition and obesity within the same communities.^16^ While the recent meta-analysis demonstrates that tuberculosis risk continues to decline across the entire BMI range, including in people with diabetes, and albeit at a slower pace among those who are overweight and obese,^15^ public health approaches must balance tuberculosis prevention goals with broader nutritional health objectives to ensure interventions do not inadvertently contribute to the growing burden of diet-related non-communicable diseases.^37^ Modelling the impact of population-level nutritional interventions beyond tuberculosis incidence to include mortality and morbidity from other diseases would help to inform this. Ultimately, further research is required to determine optimal nutritional interventions for different populations and settings. This should include participatory research methods with a broad range of stakeholders (e.g. civil society, government departments beyond health, and food system actors) to ensure interventions are locally appropriate, person-centred, and sustainable within existing systems.^38^

Our study has limitations. We applied a single risk function across all populations, though the dose-response relationship between BMI and tuberculosis risk may vary by age, sex, and setting, given varying prevalence of HIV and other potential effect modifiers. Indeed, we may potentially be underestimating the impact of eliminating undernutrition by not accounting for potential synergistic interactions between nutritional status and other leading risk factors, such as alcohol use disorders which predispose to specific nutritional deficiencies.^39^ Our analysis also relied exclusively on BMI as an indicator of nutritional status, yet other dimensions of nutrition, including micronutrient deficiencies, and dietary diversity, quality, and composition, may influence tuberculosis risk independent of BMI.^7^ Importantly, our scenarios represent idealised interventions that eliminate all BMI values below arbitrarily defined thresholds. While these scenarios are unrealistic, as with PAFs, they serve to quantify the maximum theoretical impact of potential changes in nutritional status. Finally, we did not consider the impact of eliminating undernutrition on reducing tuberculosis mortality. Given undernutrition also increases the risk of adverse outcomes among people diagnosed and treated for tuberculosis, the proportion of tuberculosis mortality averted by eliminating undernutrition may be even larger than our estimates for incidence.

In conclusion, our study demonstrates that eliminating undernutrition could prevent millions of individuals from developing tuberculosis - over a quarter of the current global adult tuberculosis incidence. The global tuberculosis response must urgently expand beyond biomedical interventions to address modifiable risk factors, which offer a critical pathway to achieving global targets that appear increasingly out of reach with current strategies. The scale-up of nutritional interventions, particularly for populations in greatest need, could have a substantial impact on the global tuberculosis epidemic while addressing a fundamental human right and delivering broader health, social, and economic benefits.

## Contributions

All authors conceived the study and critiqued the methods and approach. PJD led the analysis, with support from MJS. MJS and PJD wrote the first draft of the article. All authors contributed to the interpretation of the results, and revised and edited the article. PJD and MJS had full access to all the data in the study and had final responsibility for the decision to submit for publication.

## Data sharing

All code to reproduce this analysis is publicly available on GitHub at https://github.com/petedodd/bmitb A single archive of the public input data to reproduce this analysis has been posted on Zenodo at: https://zenodo.org/records/16900137

## Acknowledgements

MJS received funding from the NIHR UK through a Clinical Lectureship (CL-2023-16-002). CFM is also funded by BMGF (TB MAC OPP1135288, INV-059518) and NIH (R-202309-71190).

